# A gender-bias-mitigated, data-driven precision medicine system to assist in the selection of biological treatments of grade 3 and 4 knee osteoarthritis: development and preliminary validation of precisionKNEE

**DOI:** 10.1101/2021.10.06.21260506

**Authors:** Nima Heidari, James Parkin, Stefano Olgiati, Davide Meloni, Brady Fish, Ali Noorani, Mark Slevin, Leonard Azamfirei

## Abstract

**Introduction:** Osteoarthritis is a leading cause of global disability and is set to worsen with the concurrent rise in rates of obesity and an ageing population [1]. Current clinical solutions are sub-optimal with regards to their invasiveness and outcomes. Orthopaedic biologics is an emerging field that offers alternative and parallel treatment options to address this problem. Determining which patients will benefit most from these novel treatments is key in developing clinical pathways.

**Methods:** Our dataset included 329 patients treated with microfragmented fat injection (MFAT) over a 2 year period. Clinico-demographic data was recorded as well as 1-year Oxford Knee Score (OKS). The data was modelled to predict OKS 1-year response using Random Forest Regressors. Gender-bias was mitigated and outliers were hidden from the training model. The model was validated on raw test data and on a subset of patients with Kellgren-Lawrence grade 3 and 4 radiological evidence of arthritis, age greater than 64, preoperative OKS less than or equal to 27 and idiopathic aetiology of arthritis.

**Results:** The mean age and mean body mass index (BMI) of patients in our dataset was 66.4 years, 26.9 respectively. 53.5% of patients had Kellgren-Lawrence grade 4 arthritis.

The final models RMSE was 6.72, MAE was 5.38 and r-squared was 0.23 on raw test data. An RMSE of 9.77 and MAE of 7.81 was achieved when validating the model on our subset of patients. Wilcoxon signed rank tests found no evidence of predicted results being statistically significantly different to ground truth values (p ¿ 0.05).

Preoperative OKS and Kellgren-Lawrence arthritis grade was the most important feature in our model.

**Discussion:** Our model is performant and able to predict 1 year OKS response outcome within our set of patients. We have found key features of prediction and would recommend these are researched further to improve model performance.

Our dataset does not compare outcomes with other standard treatments. We also don’t compare outcomes with other biologic treatments.

Ultimately, this research can be used as a tool to benefit both patients and clinicians in a combined decision-making process.

## Introduction

Knee OA has been ranked as the 11th highest contributor to global disability with a prevalence of 3.8% (95% uncertainty interval (UI) 3.6% to 4.1%) [1]. In an increasingly ageing population with a rise in rates of obesity, Osteoarthritis is set to affect up to 15.7% of the world’s population by 2032, representing a significant cause of global disability [2]. 4.11 million (18.2%) of adults aged over 45 years in England have osteoarthritis of the knee with 6.1% being severely affected. Treating arthritis is estimated to cost the UK economy £10.2 billion in direct costs to the NHS and wider healthcare system annually. The cost of working days lost was estimated at £2.58 billion in 2017 rising to £3.43 billion by 2030. Since 2015 over 100,000 knee replacements have been performed in the UK annually [3]. Unfortunately, up to 20% of those undergoing TKA are dissatisfied with the outcome [4].

Despite the fact that patient satisfaction is an important measure of the success of a knee replacement, this is not often reported and many of the measures used to do so remain inconsistent [5]

Delaying or preventing the use of TKR would therefore be beneficial to both patients and payers [6]. The role of microfragmented adipose tissue (MFAT) in treating the pain of arthritis and improvement of function is promising; it has been safely and successfully used in the treatment of patients with early and late stages of knee OA [7], [8]. Our previous work has demonstrated the efficacy of MFAT in reducing the pain of knee OA at 1 year. We also quantified and mitigated the effect of gender bias observed in the outcomes [9] and went onto quantify the benefits achieved in a knee replacement population at 2 years [6]. The aim of this paper is to detail the process of generating an algorithm to predict the response of patients to the biological treatment of knee OA using MFAT.

## Methods

### Harvesting the Adipose Tissue and Injecting Microfragmented Adipose Tissue (MFAT)

Adipose tissue was harvested and microfragmented using previously published technique [7]. The MFAT was then injected under ultrasonographic guidance into the knee joint. The procedure was performed under sedation in an operating theatre. Following full recovery, the patients were discharged with a physiotherapy protocol.

### Dataset

The dataset used was generated from the biologic treatment of knee OA with MFAT [6], [7], [9] in 142 women and 187 men, correlating to 113980 datapoints. Variables considered were gender, age, body mass index, biological treatment type, aetiology of arthritic disease, radiological arthritic severity grade, pre-operative Oxford Knee Score and 1 year follow-up Oxford Knee Score (OKS).

Response was computed by the difference between pre-operative OKS and 1 year follow-up OKS and this was considered our outcome variable.

### Oxford Knee Score (OKS)

OKS11 comprises of 12 questions that are scored 0-4 with 0 being severe compromise and 4 being no compromise, covering pain and function of the knee. The best outcome is a score of 48 and the worst score possible is 0. This is a validated score for the measure of functional outcomes in patients undergoing knee arthroplasty [10].

All 329 patients completed questionnaires pertaining to functional and pain scores, including OKS, before treatment, and at three months, six months and one-year following treatment.

### Analysis

Analysis was performed in R version 4.1.0 using tidymodels, caret, ggdag, corrr, funModeling, rstatix, ggthemes and partykit libraries. Results were considered statistically significant at the 5% confidence level where appropriate.

### Gender-bias mitigation

Our previous work demonstrates the gender bias present within our dataset [9]. The majority class (male patients) was downsampled to match the minority class (female patients) using the caret package to address this bias. This conditional data partition package balanced our dataset whilst maintaining the observed response distribution. Our balanced data set contained 142 female patients and 144 male patients. Figure 1 demonstrates the pre and post data balancing response variable density distribution.

**Fig. 1.**
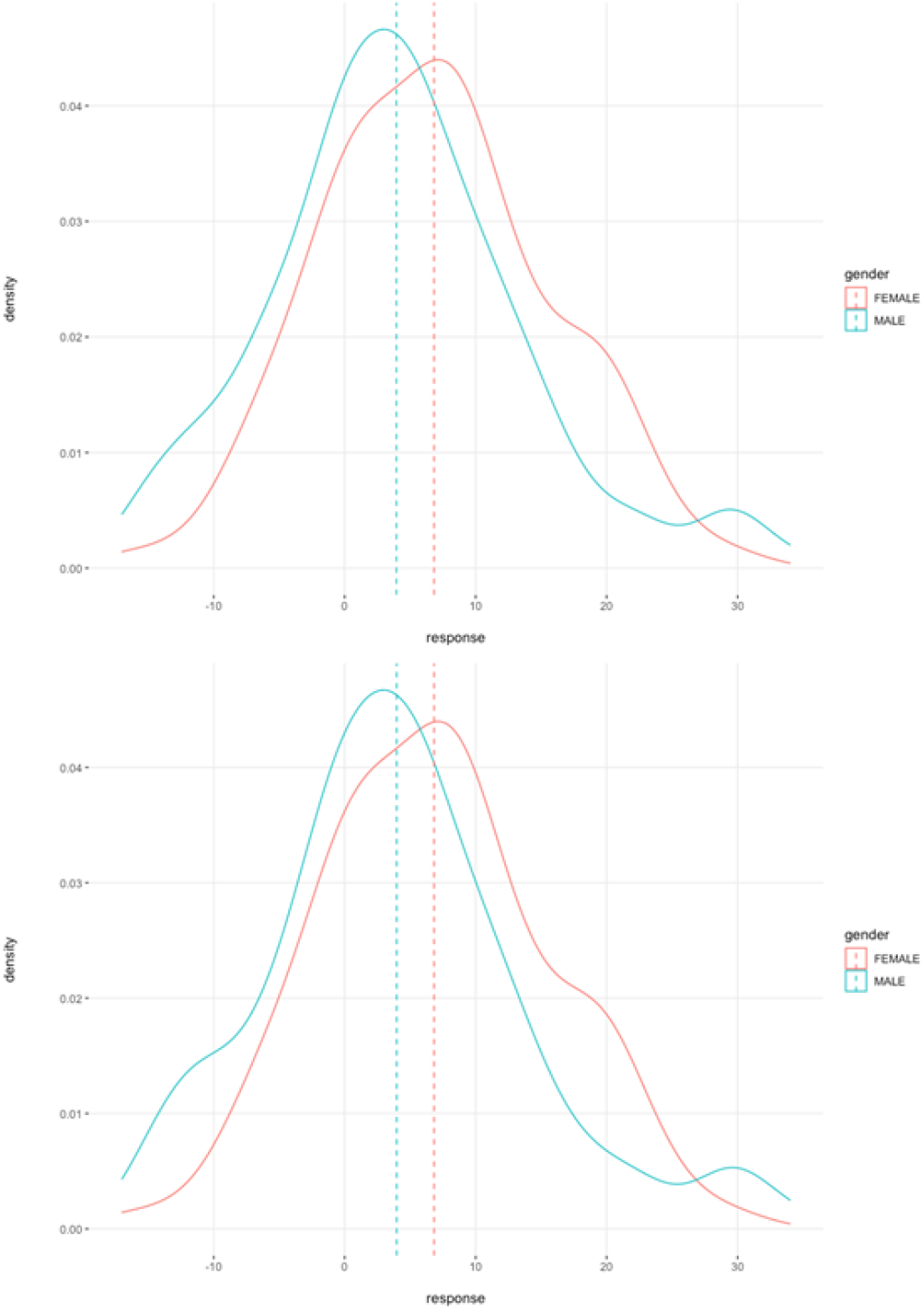
OKS response density distributions pre (top) and post (bottom) gender bias mitigation.

### Outlier detection

The dataset was further pre-processed to consider outlying response data. Two distinct training datasets were considered for modelling. Data was labelled as outlying if its response variable value fell more or less than 3 times the median absolute deviation from the median. Training dataset 1 includes outlying data (n = 231) and training dataset 2 excludes outlying data (n = 218). Both datasets were balanced as described in “Gender-bias Mitigation” before outlier detection and removal.

### Model training

All variables other than response and 1 year follow-up OKS are considered as predictor features for both model training and testing. The test set represents 20% of the size of the balanced dataset, including outliers (n = 55). Training observations were regressed against the absolute response outcome variable. Random Forest Regressors were utilised to achieve the aims of this research as they have demonstrated state-of-the-art performance in some medical fields [11], maintain some interpretability and have robust deployment solutions. Performance metrics were then produced using the test set data. Of note, the test set data included outlying observations to avoid significant data augmentation rendering the test data non-representative of the population data.

### Model validation

Models were validated using both the full test dataset and a subset of test data from patients with Kellgren-Lawrence [12] grade 3 and 4 radiological evidence of arthritis, age greater than 64, preoperative OKS less than or equal to 27 and idiopathic aetiology of arthritis. The choice of the above subset of patients is driven by the overall aim of this research. This group of patients are deemed to be suitable for TKR [9]. Thus, we aim to assess if we can mitigate or delay the need for the current suboptimal standard-of-care.

In both cases, a Wilcoxon signed rank test on paired samples between ground truth and predicted values was performed. These metrics are reported post stratification by gender.

## Results

### Descriptive analysis

Patients in our dataset had a mean age of 66.4 years on the date of procedure with a standard deviation of 10.2 years. The mean BMI of our patient cohort was 26.9 with an associated standard deviation of 4.3. As previously discussed, we had a moderate gender imbalance with 187 (56.8%) male patients and 142 (43.2%) female patients.

The majority of patients had Lipogem treatment (n = 215) and the remainder had AMPP (n = 114). 90.6% (n = 298) of patients had idiopathic arthritis followed by 6.7% (n = 22) of patients had trauma-related arthritis and finally, 2.7% (n = 9) of patients had inflammatory arthritis. 53.5% (n = 176) of patients had grade 4 radiological evidence of arthritis and 46.5% (n = 153) had grade 3 or better radiological evidence of arthritis. The mean pre-operative OKS was 31.6 with an associated standard deviation of 8.7.

Table 1 outlines our exploratory analysis stratified by gender.

**Table 1.** Overview of dataset stratified by gender

|  |  | Male | Female |
| --- | --- | --- | --- |
| Count (% of total) |  | 187 (56.8) | 142 (43.2) |
| Mean age in years (sd) |  | 65.6 (10.7) | 67.5 (9.4) |
| Mean BMI (sd) |  | 26.5 (3.6) | 27.6 (5.0) |
| Treatment (% of total) | AMPP | 56 (49.1) | 58 (50.1) |
|  | Lipogem | 131 (60.9) | 84 (39.1) |
| Aetiology (% of total) | Idiopathic | 164 (55.0) | 134 (45.0) |
|  | Inflammatory | 6 (66.7) | 3 (33.3) |
|  | Traumatic | 17 (77.3) | 5 (22.7) |
| OA Grade (% of total) | 0 | 0 (0.0) | 1 (100.0) |
|  | 1 | 14 (77.8) | 4 (22.2) |
|  | 2 | 25 (42.4) | 34 (57.6) |
|  | 3 | 43 (57.3) | 32 (42.7) |
|  | 4 | 105 (59.7) | 71 (40.3) |
| Mean pre-op OKS (sd) |  | 33.1 (8.8) | 29.7 (8.1) |

The Pearson correlation coefficient was calculated for each continuous feature against every other continuous feature.

Figure 2 visualises this relationship in the form of a correlation matrix.

**Fig. 2.**
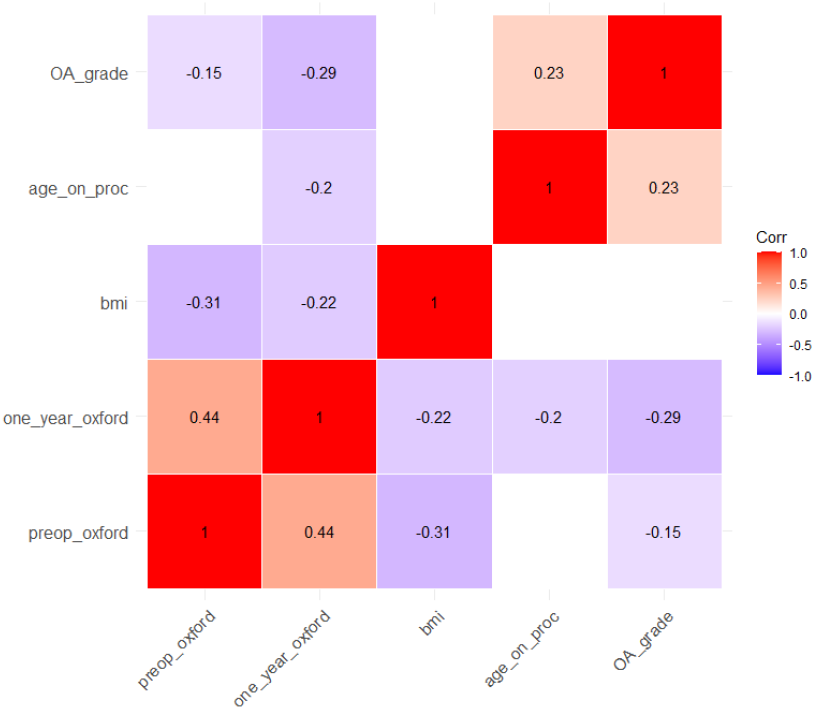
Correlation matrix showing the Pearson correlation coefficients for continuous features. Statistically insignificant coefficients have been blanked out.

We note that the strongest statistically significant linear correlation observed was between pre-operative OKS and one-year OKS. BMI and radiological grade of arthritis were negatively linearly associated with pre-operative OKS at values of −0.31 and −0.15 respectively. Age at procedure, BMI and radiological grade of arthritis were all negatively associated with OKS at one year. We observed a 29.0% reduction in strength of negative associated to BMI between pre-operative OKS and one-year OKS. Conversely, we observed a 93.3% increase in the strength of negative association to radiological grade of arthritis between pre-operative OKS and one-year OKS.

### Model results

Our model was trained without outliers as outlined in “Outlier detection”. Training OOB prediction error (MSE) was 50.9 with an associated r-squared value of 0.22.

The model was tested on data without outlier removal. Test RMSE was 6.72, MAE 5.38 and r-squared was 0.23. Figure 3 demonstrates the actual vs predicted results of our final model, stratified by gender.

**Fig. 3.**
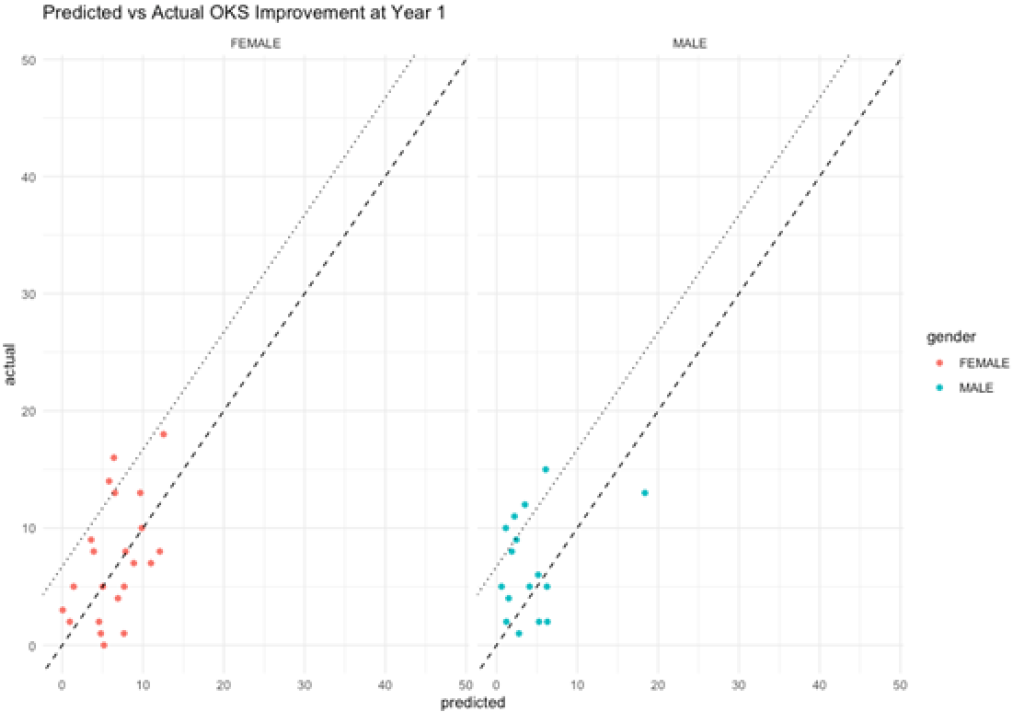
Predicted vs. actual values stratified by gender with no filtering on the test set.

The final model’s predictions were compared with ground truths using Wilcoxon signed rank tests. Male and female gender stratified predictions had p-values of 0.93 and 0.92 respectively, providing us with no evidence of statistically significant differences in sample medians between ground truth and predicted values. Figure 4 visualises this result.

**Fig. 4.**
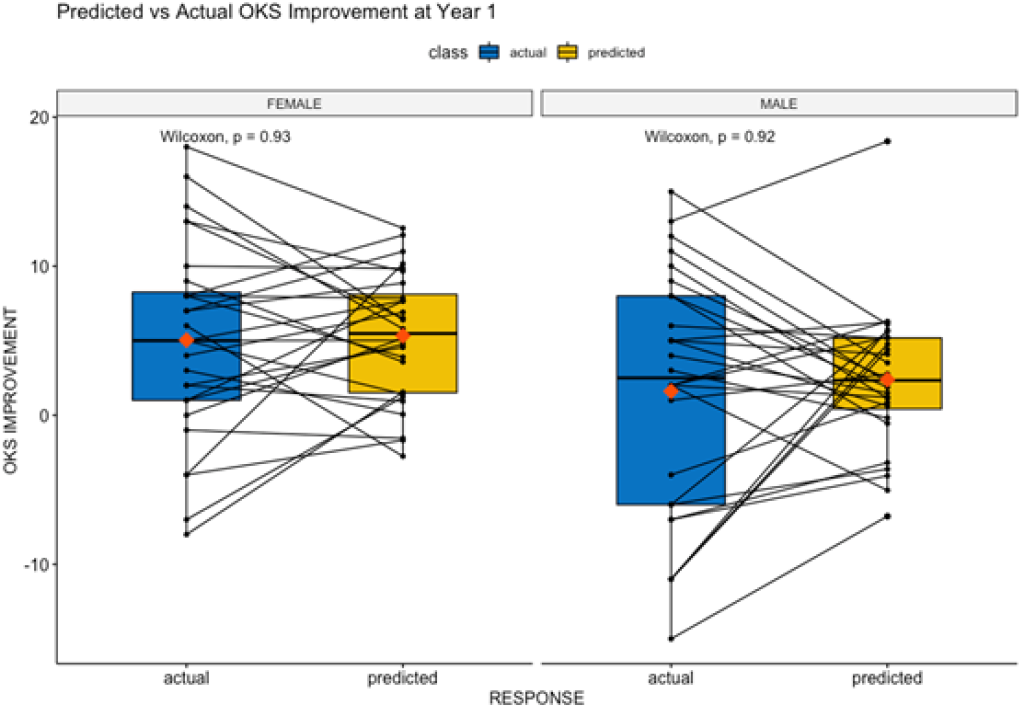
Predicted vs. actual OKS values: displaying median values and Wilcoxon significance levels.

### Model validation

Further testing of the model using patients suitable for TKR (as outlined in methods) demonstrated a RMSE of 9.77 and MAE of 7.81. Figure 5 demonstrates the actual vs predicted results of our final model using this subset data for testing.

**Fig. 5.**
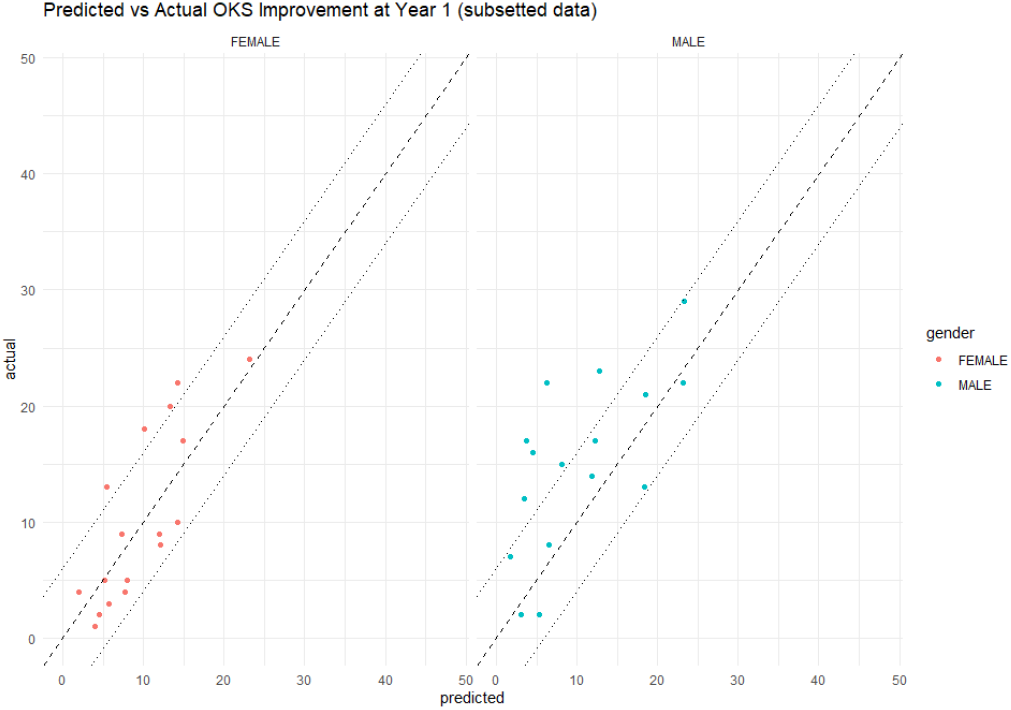
Predicted vs. actual OKS values filtered by suitability for TKR (as described in “Methods: Model validation”).

### Model Interpretation

To interpret and explain our results, a conditional inference tree was computed outlining the most significant features our models identified in order to achieve performant results. Figure 6 demonstrates three major features contributing to an observation being classified into 1 of 5 response groups.

**Fig. 6.**
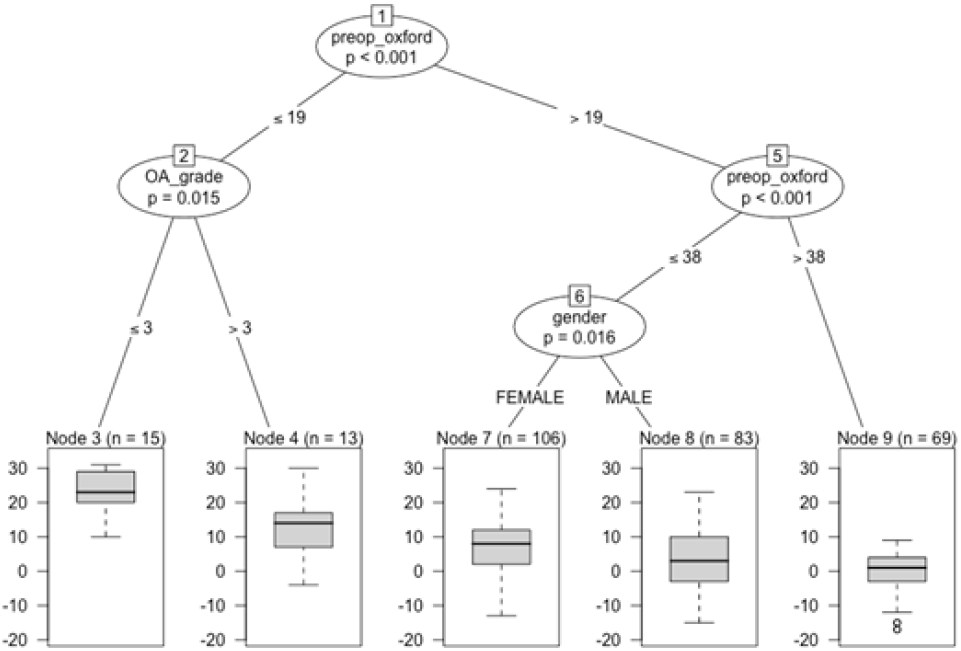
Conditional inference tree computed using the final trained model.

- We find that preoperative OKS was the most important feature and takes its place at the beginning of the tree. It further divides patients with an OKS of more than 19 into 2 lower responding groups.
- Radiological arthritic severity grade subdivides a group of highly responsive patients into certain and less certain response groups.
- Even after gender bias mitigation, we find gender to be a key feature in refining uncertain responders.

## Discussion

Medicine is one of the last disciplines to truly embrace the data-driven approach. Many of the surgical treatments in orthopaedics are only now being supported by a data-driven approach. To address the lack of data-driven decision making in medicine, we must have the engagement of clinicians and patients as well as the payers to arrive at decision that are both clinically robust and economically sound.

All our models were performant, interpretable and deployable. We have determined key features pertaining to accurate prediction of response to treatment and can therefore make suggestions regarding future research.

The use of the precisionKNEE clinical decision making tool requires external validation with an appropriate study. We believe it needs to be viewed as a diagnostic tool, much like imaging, providing additional information for the clinician and the patient to navigate the complex array of treatments available. Further studies are required to assess precisionKNEE in a clinical setting. Such a study should incorporate more resolute features representing 2 or 3-dimensional imaging as our research indicates these are important in determining response to biological treatment. The SPIRIT-AI and CONSORT-AI initiative provide guidance to improve the transparency and completeness of reporting of clinical trials evaluating interventions involving artificial intelligence as such any study needs to be designed with these in mind.

### Limitations

Despite being one of the largest datasets of its kind, upon stratification we observe groups of patients with minimal observations and thus we cannot conclude external validity in the findings pertaining to these groups.

Kellgren-Lawrence classification of arthritis was found to be an important feature during modelling. This measure is crude and its relationship to patient symptons remains.

MFAT is the only treatment used in our model. With inclusion of other modalities of treatment such as other biologics including BMAC, PRP and n-Stride as well as TKR a true clinical decision making support tool can be developed.

This may be viewed by clinicians as an incursion into their territory of decision making. However, patients find these very useful and by providing a personalised prediction of the possible outcome, they will be able to play a more meaningful role in the decision-making process.

## Conclusions

By using a validated dataset for the use of MFAT for the treatment of Knee OA, we have built an accurate model for the prediction of response. This model has been validated in a general test set representative of the patients eligible for the treatment. We further validated this model in a subset of patients who are suitable for TKR and thus could potentially benefit from biologic treatment and mitigate the risk of the surgical intervention.

This tool is of benefit to patients and clinicians in a combined decision making process to choose the best treatment.

Continued data collection in the form of registries (such as this dataset) will allow for tuning of the model and mitigating bias generated through under-representation of a particular group.

## Data Availability

Data used for the production of this manuscript is not publicly available.

## Author contributions statement

S.O. and N.H. were responsible for conceptualization; S.O., J.P., N.H. were responsible for methodology; S.O., J.P. and D.M. were responsible for the software; S.O., J.P., and N.H. were responsible for the formal analysis; N.H. and A.N. were responsible for the investigation; B.F. was responsible for the resources; N.H. was responsible for data curation; J.P. and N.H. were responsible for writing—original draft preparation; N.H. and J.P. were responsible for writing—review and editing; N.H., S.O. and J.P .were responsible for visualization; N.H. and S.O. were responsible for supervision; B.F. was responsible for project administration. All authors have read and agreed to the published version of the manuscript. S.O., N.H. and J.P are responsible for the algorithms and processes on bias mitigation.

## Ethics Statement

This study was carried out in compliance with the rules of the Helsinki Declaration and International Ethical Regulations, including all subsequent amendments, under the approval of the Research Ethics Committee of the “George Emil Palade” University of Medicine, Pharmacy, Science and Technology of Targu Mures, Romania (research approval number 1464/2021).

## Acknowledgments

The authors acknowledge Angela Cullen as the Data Manager and Analyst.

